# Impact of Indolent Schistosomiasis on Morbidity and Mortality from Respiratory Tract Infections in Preschool Age Children from a Schistosomiasis Endemic Area

**DOI:** 10.1101/2020.11.06.20227173

**Authors:** Tariro L. Mduluza-Jokonya, Arthur Vengesai, Luxwell Jokonya, Amanda Thakataka, Herald Midzi, Takafira Mduluza, Elopy Sibanda, Thajasvarie Naicker

## Abstract

**Introduction:** Pneumonia is the biggest child killer, after the neonatal period. This is especially so in children from developing countries who are exposed to other infections simultaneously. In this article we investigated the impact of indolent *Schistosoma haematobium* infection on background of a respiratory infection

**Method:** A cross sectional study with 237 preschool age children with a respiratory infection, was performed during winter months in a schistosomiasis endemic area. Participants were clinically examined and investigated appropriately. Upper respiratory tract infection (URTI) and pneumonia were defined and classified as per IMCI and WHO guidelines, respectively. *S. haematobium* infection diagnosis was by urine filtration on urine collected over three consecutive days. Data was analysed using SPSS.

**Results:** *S haematobium* infection prevalence was 29% (69). Prevalence of repiratory infections were as follows: common cold 79% (188), pneumonia 15% (36) and severe pneumonia 6% (15). Eighty-one percent of participants with the common cold were *S. haematobium* negative, whilst 80 % of those with severe pneumonia were infected. Schistosomiasis infected children were at greater odds of developing; pneumonia (aOR=3.61 (95% CI 1.73-7.55) and severe pneumonia (aOR=21.13 (95% CI 4.65-95.89). High intensity S. haematobium infection was associated with an increased risk of severe pneumonia RR= 23.78(95% CI 6.86-82.32). Mortality from coinfection emanated from severe pneumonia and severe *S. haematobium* infection intensity (RR= 26.56 (95% CI 1.49 to 473.89). Number needed to harm (NNH) for *S. haematobium* infected children who develop respiratory tract infection was 4:1 for pneumonia and 5:1 for severe pneumonia.

**Conclusion:** The study demonstrated that coinfection with Schistosomiasis increases morbidity and mortality from respiratory tract infections by up to 20 times in children less than five years old. There is need to cover schistosomiasis screening and treatment in children under 5 years old to avert mortality and morbidity due to coinfection with respiratory infections.

## Background

Pneumonia, an infection that inflames the respiratory system air sacs causing cough, fever and difficulty to breath, was described over 2400 years ago by Hippocrates (1). Despite modern day medicine, it is still the leading cause of death world-wide (2). It is the biggest child killer, estimated to have 150.7 million new cases per year, of which 11-20 million are severe enough to require hospital admission (3). In Zimbabwe, pneumonia is the leading cause of childhood deaths after the neonatal period (4).

Pneumonia frequently starts as an upper respiratory tract infection which progresses to the lower respiratory tract (5). Micro-aspiration of contaminated secretions infects the lower airways to cause pneumonia (6). Pneumonia is at times described as a bacterial infection of the lung that require antimicrobial treatment (1). Though the pathogenic organism can be viruses, fungi or parasites (7). Common implicated agents include: *streptococci pneumoniae, Haemophilus influenzae, Chlamydophila pneumoniae*, rhinoviruses, coronaviruses, influenza virus, parainfluenza virus, adenovirus and respiratory syncytial virus (5). About 45% of cases in children are due to mixed infections of viruses and bacteria (8). A causative agent may not be isolated in 50% of the cases despite careful testing (9). Ninety-five percent of clinical cases in young children occur in developing countries (10). In developing countries, there are endemic conditions that cause harm to the lungs. Damaged lungs are more susceptible to pneumonia acquisition (11).

Schistosomiasis is common in Zimbabwe (12). It is caused by a trematode (worm) of the genus *Schistosoma* (13). There are 24 species in the genus, seven can infest humans, in Zimbabwe the two common species are *S. haematobium* and *S. mansoni*, with the former having a higher prevalence (12,14 -18). *S. haematobium* is commonly associated with genitourinary symptoms, however the infestation process affects other organs, such as the lungs (13), especially during the worm developmental stages. Pulmonary schistosomiasis has been described in two forms, acute pulmonary schistosomiasis (APS) and chronic pulmonary schistosomiasis (CPS) (19, 20). APS is reported in naï ve travelers who acquire schistosomiasis and CPS being described as pulmonary hypertension common in the *S. mansoni* species (21, 22).

Indolent in medical terminology refers to pathology causing little or no pain; inactive or relatively benign (23). In most Preschool age children (PSAC) there is *S*.*haematobium* infection with no visible signs, this is the indolent phase of schistosomiasis. As previously reported, children are exposed *in utero*, and are again exposed postnatally (24). In most PSAC, symptoms begin when children are of the school-going age (15). The pulmonary effect of S. haematobium infection has not been described in PSAC. Despite being the second most common parasitic infection in the world, infecting about 50 million PSAC, the impact of the coinfection of schistosomiasis and upper respiratory tract infections have not been described, in endemic countries (25, 26). In this study we investigated the association between *S. haematobium* infection and pneumonia in children presenting with symptoms of respiratory tract infection under the age of five in a district in Zimbabwe.

## Methodology

### Study site and design

The study was a cross-sectional study, performed during winter months. It was carried out in the Shamva district (31°40′0” E longitude and 17°10′0” S latitude) in Mashonaland Central province, Zimbabwe (27). It lies 945 m above sea level, the climate is warm and temperament with an average temperature of 20.2 °C and an annual rainfall of 887 mm. Located in the Mazowe valley, Shamva district is an area with high farming activity due to its fertile soil. Residents get their water supply from Mazowe river which spans through the district and is a huge source of alluvial gold which the vast majority of population survives through panning (27). Shamva District is a *S. haematobium* infection endemic area with the highest prevalence of schistosomiasis in Zimbabwe at 62.3%, in PSAC schistosomiasis prevalence was recorded to be 10% (28).

### Sample size

Children aged five years and below with upper respiratory tract infection were recruited into the study. Mothers were requested to bring their children to the clinic or to Expanded Programme on Immunization (EPI) meeting points. The required sample size was calculated to be 138 participants using the formula below with *S. haematobium* prevalence of the area recorded as 10% (15):

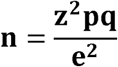

Where **z** is the ***z*** value for the 95% confidence interval, that is alpha□=□5% (*z*□=□1.96)

p□=□proportion/prevalence of the outcome to be investigated (*p*□=□0.10)

q□=□1-*p*□=□0.90

e =□precision for the given confidence interval expected expressed as a decimal (*e*□=□0.05)

*n* **=** 138

Using simple random sampling in the Shamva village, we recruited a total of 237 study participants.

### Study inclusion criteria

Children under five years old with signs and symptoms of an upper respiratory tract infection were recruited into the study after exclusion of common non schistosomiasis co-morbidities in the area which included malaria, tuberculosis, HIV infection, malnutrition, asthma, allergies and family history of atopy.

### Ethical statement

Ethical approval was obtained from Medical Research Council of Zimbabwe (MRCZ/B/1854). Gatekeeper approval was obtained from the Provincial and District Medical Directors and Community Leaders. Informed consent was obtained from the parent/guardian of the children. All participants with confirmed disease were offered treatment and parents/caregivers were counselled. Participants with severe pneumonia signs were immediately transferred to the nearest health facility for further management.

### Clinical examination

The clinical examination was conducted on PSAC (n = 237) by **two** medical practitioners independent of each other at health facilities. Each clinician first took the history and then examined each child according to the standard protocol (29,30) with special attention to the respiratory system.

### Respiratory System Examination

Inspection: Cyanosis of lips and nails, clubbing of fingers, respiratory rate, depth and character of respiration, chest symmetry and expansion, use of accessory muscles, retractions Palpation: Tenderness, subcutaneous crepitation, position of trachea, degree of chest expansion (in cm with tape), tactile fremitus Percussion: percussion notes (resonance, hyper-resonance, dull, flat), diaphragmatic excursion Auscultation: Character of breath sounds (vesicular, bronchovesicular, bronchial, tracheal), crackles, wheezing, friction rub, vocal resonance

### Upper Respiratory Tract Infection (URTI) Diagnosis

URTI was diagnosed on clinical signs and symptoms after excluding allergy and influenza according to IMCI guidelines. Signs and symptoms clinicians used to make a diagnosis included sneezing, coughing, sore throat (reluctance to feed/unable to feed), nasal discharge and nasal congestion (9,31–33).

### Pneumonia Diagnosis

Pneumonia was defined as per WHO guidelines (7, 34).

Class 1: No pneumonia (cough/cold) - Normal respiratory rate, no use of accessory muscles for breathing.

Class 2: Pneumonia - Tachypnoea: >50 breaths per minute (infants 2-11months), >40 breaths per minute (children 12-59 months), No use of breathing accessory muscles.

Class 3: Severe pneumonia - Use of breathing accessory muscles with or without tachypnoea, Any general danger signs: unable to drink, drowsiness, convulsions, reduced level of consciousness, Oxygen saturation SPO_2_ < 90%, Stridor in a calm child.

### *S. haematobium* Infection Diagnosis

Urine samples were collected by giving the caregiver an open-mouth urine container, children 1 year and below used paediatric urine collectors attached by a clinician. The samples were examined for macrohematuria using the Uristix reagent strips (Uripath, Plasmatec, UK) dipped into fresh, well-mixed urine for 40□sec and the test area was compared with a standard colour chart as per manufacturer’s instructions. Approximately 50 ml of urine sample was collected from each participant on three consecutive days. The samples were collected between 10am and 2pm, processed within two hours of collection by the urine filtration method and were examined using microscopy for *S. haematobium* eggs detection, as previously described (35). The number of eggs were reported per 10ml of urine. The parasitology team recorded the results and the clinical team was blinded from them.

### Tuberculin Test for Tuberculosis

About 0.1ml of liquid containing five tuberculin units purified protein derivative was injected into the epidermis of the forearm. The result was read after 48hrs, an induration of >5mm was read as positive and the child would be excluded from the study. Nonetheless, the caregivers were referred to the nearest health care facility and the village health worker and community nurse followed-up (31).

### Rapid Diagnostic Tests

Rapid diagnostic tests were done for HIV infection and malaria with the caregiver’s consent. Children exposed to HIV were excluded from the study, and they received counselling on the importance of having the child tested after weaning.

### Malnutrition Diagnosis

Height and weight were measured with the participants in light/no clothing, an infantometer baby board was used to measure height and for weight we used a baby scale. MUAC: measurement was done on the left arm mid-point between the shoulder and the elbow tip, with the arm relaxed and hanging down the body. Height and weight for age charts were used to assess nutritional status. The caregivers were counseled concerning the correct nutritional requirements and the importance of having a balanced diet. Children with malnutrition were referred to the local health facility for further management.

### Statistical Methods

Data analysis was performed using IBM SPSS Statistics version 23. Initial analysis was to determine an association between participants with URTI advancing to pneumonia and *S. haematobium* infection status. The statistical methods applied were the descriptive statistics, bivariate and multivariate logistic regression modelling. The multivariate logistic regression models were fitted to adjust for potential confounding factors for the five manifestations with three explanatory variables, that is, sex, age and schistosomiasis infection. The effect of different factors on the prevalence of schistosome infection and morbidity was determined using logistic regression and the results reported as adjusted ORs (AORs) and 95% CI, along with the test for significance, as previously described (36). Second analysis was to determine the effects of different intensities of *S. haematobium* infection as a risk factor for developing pneumonia or severe pneumonia. We calculated the population attributable fraction (PAF) of the *S. haematobium* infected participants with different infection intensity having pneumonia using the formula:

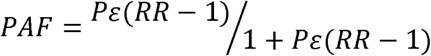

Where P_e_ = Percentage of population exposed, RR=relative risk

Final analysis was to determine the impact to the population of *S. haematobium* infection as a risk factor for pneumonia. For this we used the population attributable risk (PAR) of getting pneumonia in the presence of *S. haematobium* infection calculated using the formula (37,38):

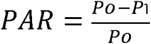

Where P_o_ = participant with the condition divided by total participants

P_1_ = number with condition unexposed divided by total participants unexposed

Number needed to harm (NNH) in the study population was calculated using the following formula:

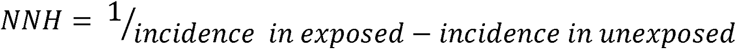

*S. haematobium* infection was defined as the arithmetic mean egg count/10mL of at least two urine samples collected on three consecutive days.

## Results

### Demographics

A total of 319 children under five years from 19 Shamva district villages were screened. (Figure 1). Two hundred and thirty-seven preschool age children with signs and symptoms of URTI were recruited into the study and followed up. The number of males was 129 (54%) and females 108 (46%). The age was normally distributed with mean (SD) age of 3.2 (1.2). *S. haematobium* prevalence was 29% (69) (Table 1). Amongst the study participant, 84% (201) had exposure to contaminated water, whilst 15% reported no exposure to contaminated water.

**Table 1:** Characteristics of the Study Participants

| Characteristic |  | Total (N=237) |
| --- | --- | --- |
| Age mean (SD) |  | 3.2 (1.2) |
| Gender |  |  |
| Males |  | 54% (129) |
| Females |  | 46% (108) |
| Pneumonia class among participants |  |  |
| No pneumonia (Common cold ) |  | 182 (77%) |
| Pneumonia |  | 37(16%) |
| Severe pneumonia |  | 16(7%) |
| Risk Factors |  |  |
| Exposure to contaminated water | No | 15% (36) |
|  | Yes | 84% (201) |
| Indoor pollution | No | 5.5%(13) |
|  | Yes | 94.5%(224) |

**Figure 1:**
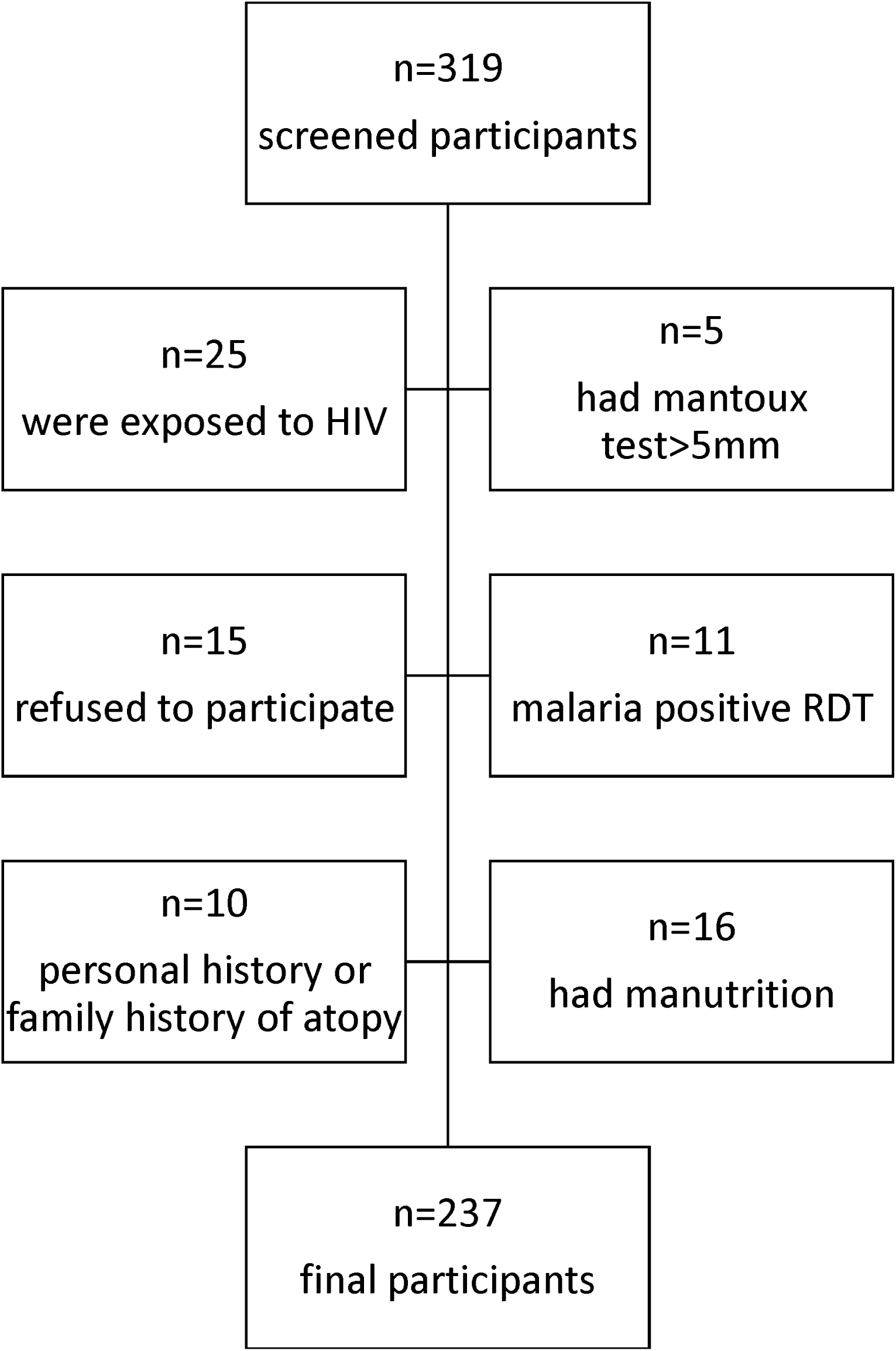
Profile showing the inclusion and excluded of the children and the final participants included in the study.

### Relationship between *S. haematobium* Infection and Pneumonia

After examination, 79% (182) of children had a simple cold or cough, 15% (36) had pneumonia, 6.3 % (15) had severe pneumonia and 33% (5) of the severe pneumonia participants deceased (Table 2). Eighty one percent of participants with the common cold were *S. haematobium* negative, whilst 80 % of participants with severe pneumonia were *S. haematobium* positive and 57% of *S. haematobium* positive had pneumonia.

**Table 2:** Relationship between *S. haematobium* Infection and Pneumonia

|  |  | No Pneumonia<br>(common cold)<br>N=182 | Pneumonia<br>N=37 | Severe<br>pneumonia<br>N=16 | Mortalities<br>N=5 |
| --- | --- | --- | --- | --- | --- |
| *S. haematobium*<br>Infection status | negative | 81% (148) | 43%(16) | 12% (2) | 0 |
|  | positive | 19% (34) | 57%(21) | 88%(14) | 5 |
| Total prevalence % (n) |  | 79%(182) | 15% (37) | 7% (16) | 2% (5) |
| adjusted odds ratio (95% CI) |  | **0.16(0.08-0.33)\*** | **4.16(2.01-8.59)\*** | **21.13(4.65-95.89)\*** | **9.79( 3.82- 17.53)\*<sup>a</sup>** |
| Relative risk (95% CI) |  | **0.26(0.16-0.43)\*** | **3.20(1.78-5.75)\*** | **17.04(3.98-73.01)\*** | **26.56( 1.49-473.80)\*<sup>a</sup>** |
| Population attributable<br>fraction |  | - | 31.61% | 82.38% | 27.88% |
| Mortality |  | 0% | 0% | 31%(5) |  |

\* significant at 5% level of significance( $p < 0.05$ , <sup>a</sup> in relation to severe pneumonia

The study participants after adjusting for sex and age, *S. haematobium* infection status and indoor pollution, the infected children were at higher odds of presenting with the following: pneumonia (aOR = 3.61 95 % CI 1.73-7.55) and severe pneumonia (aOR =21.13(4.65-95.89) and mortality (aOR = 9.79 95% CI 3.82-17.53). The risk of having pneumonia or severe pneumonia in the *S. haematobium* infected individuals increased (Figure 2), with the following relative risk (RR) noted: Pneumonia (RR=3.20(95% CI 1.78-5.75) and severe pneumonia (RR=17.04 (3.98-73.01) with a mortality (RR = 26.56 (95% CI 1.49-473.80). The PAF of pneumonia in schistosomiasis positive children was 31.61% and for severe pneumonia was 82.38%. PAF due to respiratory tract and *S. haematobium* co-infection was 28%.

**Figure 2:**
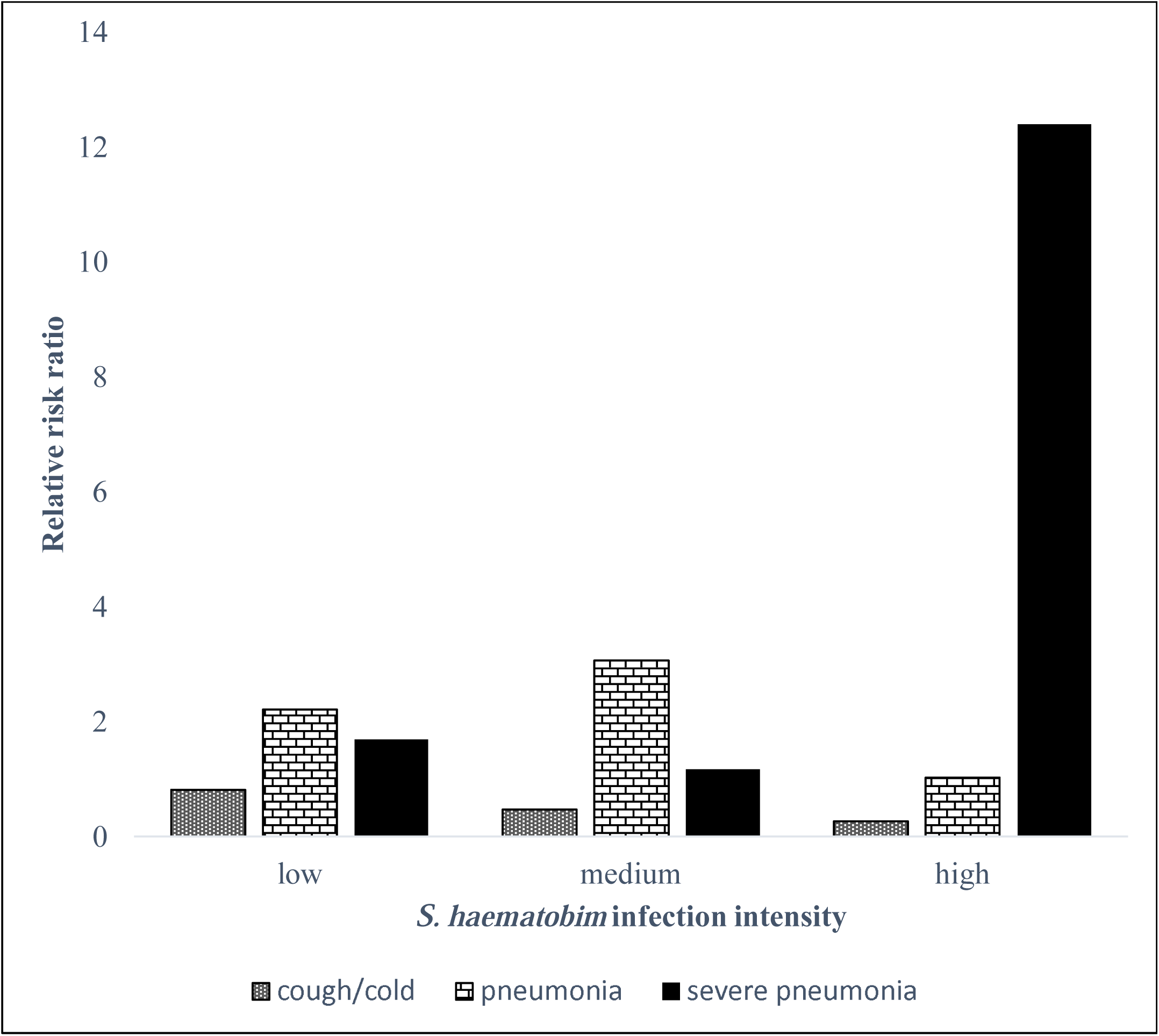
Relative risk ratio of *S. haematobium* infection intensity measured by eggs in 10mls of urine and pneumonia diagnosed by clinical examination.

### *S. haematobium* Infection Intensity Association with Respiratory Infection

Children with low intensity *S. haematobium* infection were at a higher risk of developing pneumonia with RR=(2.62 (95% CI 1.13-6.08) (Figure 2). *S. haematobium* infected participants with medium infection intensity were also more likely to present with pneumonia (RR=4.31 (95% CI 1.45-12.81). High infection intensity was associated with a higher risk of having severe pneumonia (RR=23.78 (95% CI 6.86-82.32). Having high intensity infection had a RR=(12.22 (95% CI 1.86-80.10) of dying. In terms of population attributable fraction in low *S. haematobium* infection intensity status attributed 26.79% of pneumonia, mild infection mainly attributed to pneumonia PAF=16%. In high infection intensity had a high PAF with severe pneumonia of 42% (Figure 3).

**Figure 3:**
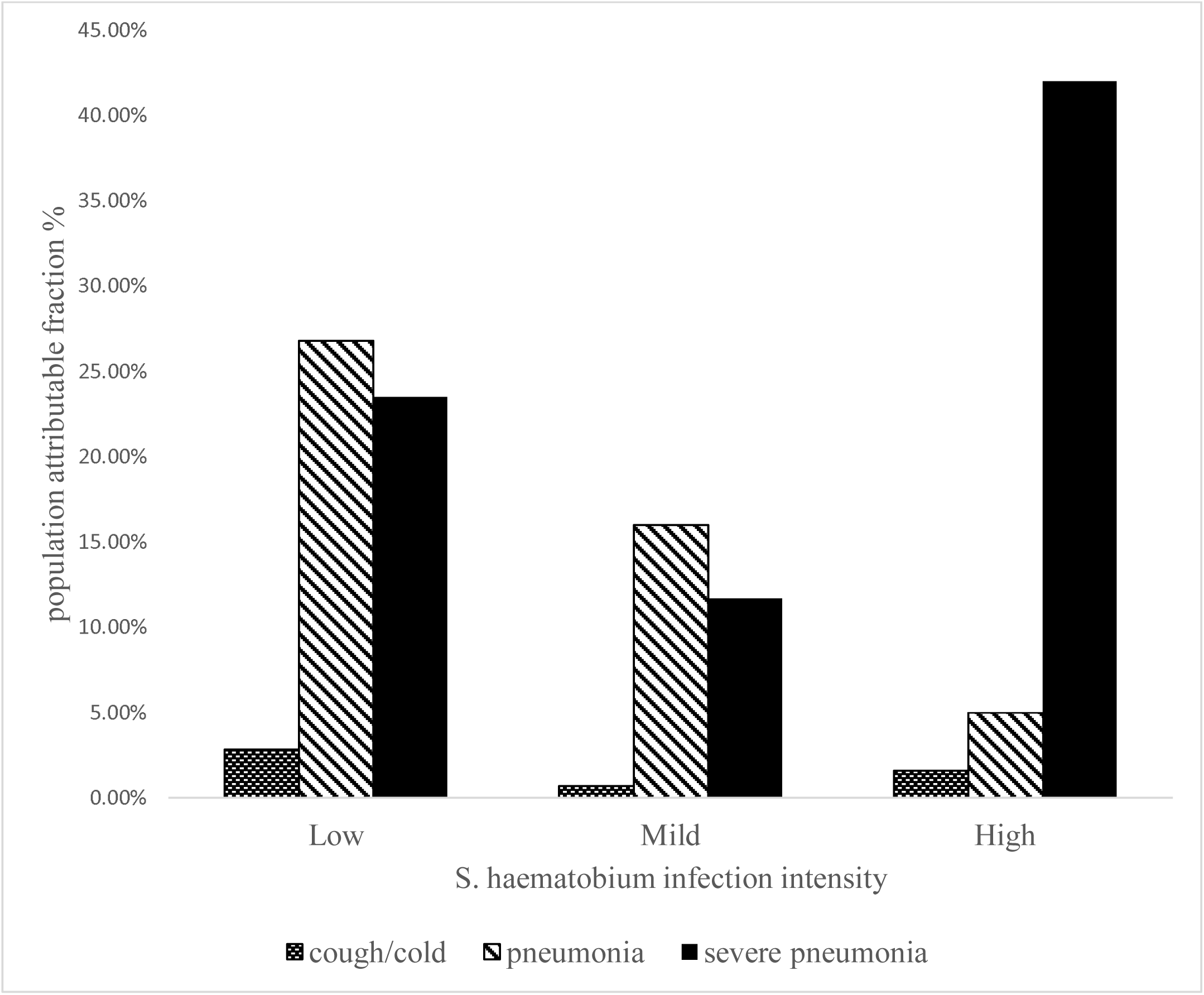
*S. haematobium* infection intensity and population attributable fraction

### Population Impact of *S. haematobium* Infection on Development of Pneumonia

Population attributable risk percentages of schistosomiasis to pneumonia were as follows: 39.1% for pneumonia and 85.7% for severe pneumonia. Number needed to harm (NNH) by not treating *S. haematobium* infection in the Shamva district was as follows:

1. For every **four** children exposed **one** will get pneumonia
2. For every **five** children exposed to *S. haematobium* infection **one** will get severe pneumonia in the event of acquiring an upper respiratory tract infection.

## Discussion

This study reveals that there is a strong association between having *S. haematobium* infection and risk of pneumonia or severe pneumonia. In general, more than half of the severe disease burden was attributable to *S. haematobium* infection. We also report an association between high infection intensity status and development of severe pneumonia, whilst low and mild infection intensity were more associated with URTI progression to pneumonia. Participants who deceased all had severe pneumonia. Preschool age children with indolent schistosomiasis who catch the common cold were at a higher risk of acquiring pneumonia or severe pneumonia, according to our findings. This could be due to lung damage that occur during *S. haematobium* infection predisposing the children to severe disease progression (20). Pulmonary schistosomiasis occurs as early as the first exposure to infection, chest x-rays of individuals with acute schistosomiasis have shown widespread non-specific infiltrates which were accompanied by respiratory symptoms which include wheezes and dyspnea (39). These respiratory signs and symptoms were reported to last for months after initial harm occurs (19). Despite resolution of wheezes, the infected person is still exposed to further lung injury during the chronic schistosomiasis phase (40–42).

This study shows that infection intensity was associated with URTI development turning into the severe form of lower respiratory infection. Children with high intensity *S. haematobium* infection had a significantly increased risk of severe pneumonia and death. This is in keeping with previous reports on schistosomiasis and morbidity, which showed increased morbidity with increased infection intensity (43–45). Schistosome eggs trapped in the host tissue are the major cause of morbidity (46–48). In chronic schistosomiasis, eggs of the parasite are released into the host human and are spread throughout the body and mainly into the lung where they cause pulmonary arteritis (46). Infection intensity describes the parasitic load and is measured as the number of eggs excreted, thus the higher the infection intensity the more the associated morbidity.

To our knowledge, this is the first study to report a possible exacerbating effect of *S. haematobium* infection on respiratory tract infection. We report *S. haematobium* infection to have a significant population attributable fraction to severe pneumonia of 82.35% in our study population. Furthermore, we found that for every four children with *S. haematobium* infection who acquire an URTI, one will develop pneumonia. Whilst for every five *S. haematobium* infected PSAC who acquire an URTI, one will develop severe pneumonia. There have been reports of exacerbation of other clinical conditions with schistosomiasis, like typhoid and malaria (49 – 52). To our knowledge, no reports have looked at the effect of schistosomiasis on respiratory tract infections, particularly in PSAC.

Our study is of significant public health importance, notably in Zimbabwe there are about 1324 hospital admissions in PSAC due to pneumonia and severe pneumonia every month (49). Despite strides in improving mortality by tackling malnutrition, HIV infection and the introduction of polymer vaccines, the numbers still remain high. Thus, including routine screening and early treatment of *S. haematobium* infection in a schistosomiasis endemic area may have immense benefits in preventing severe pneumonia. There is need for policy makers in endemic areas to further strengthen the drive to screen and treat schistosomiasis in PSAC in a bid to decrease the mortality and morbidity associated with respiratory infections. Furthermore, in the case of a respiratory pandemic, like SARS coronavirus-2, the impact on children under five in schistosomiasis endemic area has the potential to be deadly.

Limitations to our study was the fact that it was carried out during a low schistosomiasis transmission season, although it was a high URTI season. Our second limitation was that the type of pathogen which caused the URTI could not be identified. Study strengths include the fact that our sample size was more than the calculated size and the study was done in a high schistosomiasis endemic zone, thus our results may be related to other endemic areas. This study is of great importance in the public health sector, since respiratory tract infections are the leading cause of children under-five years, mortality after the neonatal period. Eliminating schistosomiasis could mean improving children under-five years mortality in endemic areas.

## Conclusion

This study demonstrates a harmful association between *S. haematobium* infection and respiratory tract infections. *S. haematobium* infected children displayed a significantly increased risk of developing pneumonia and severe pneumonia. Furthermore, the number needed to harm in our study population was significantly high. This emphasizes the immense benefit of early diagnosis and treatment of *S. haematobium* infection in PSAC, especially during a global pandemic of a respiratory disease.

## Data Availability

The statistical data on the parasitology and clinical scores used to support the findings of this study are available from the corresponding author upon request.

## Conflict of Interest

The authors declare that there is no conflict of interest.

## Funding Sources

The study received funding from the TIBA. This research was commissioned by the National Institute of Health Research (NIHR), Global Health Research Programme (16/136/33) using UK aid from UK Government. The views expressed in this publication are those of the authors and not necessarily those of the NIHR or the Department of Health and Social Care.

## Acknowledgements

We would like to acknowledge the Ministry of Health and Child Care, the Medical Research Council of Zimbabwe, village health workers, nursing staff, parents and children from Shamva. A special thanks to members of the Biochemistry Department at the University of Zimbabwe for technical support during field parasitology and sampling. Our most profound gratitude to the participants and their parents or guardians for taking part in this study.

## Author Contributions

TLMJ, LJ, TN, ES and TM conceived and designed the study. TLMJ, LJ, AT, AV, HM, ES and TM performed the clinical examination or parasitology and the data analysis. TLMJ wrote the first draft and all authors contributed to the manuscript and revised the final version.

## Notes

### Competing Interest Statement

The authors have declared no competing interest.

## References

1. Tsoucalas G, Sgantzos M. General Medicine□: Open access Hippocrates, on the Infection of the Lower Respiratory Tract among the General Population in Ancient Greece. 2016;4(5).

2. Loubet P, Tubiana S, Claessens YE, Epelboin L, Ficko C, Bel J Le. Community-acquired pneumonia in the emergency department□: an algorithm to facilitate diagnosis and guide chest CT scan indication. 2019.

3. Murray CJL, Vos T, Lozano R, Naghavi M, Flaxman AD, Michaud C, et al. Disability-adjusted life years (DALYs) for 291 diseases and injuries in 21 regions, 1990 – 2010□: a systematic analysis for the Global Burden of Disease Study 2010. 2014;1990–2010.

4. Zimbabwe national statistics survey. Demographic and health surveys.

5. Drain PK. Acute Respiratory Diseases and Pneumonias. 2011.

6. Tang J, Chen J, He T, Jiang Z, Zhou J, Hu B, et al. Diversity of upper respiratory tract infections and prevalence of Streptococcus pneumoniae colonization among patients with fever and flu-like symptoms. 2019;1–10.

7. WHO. Revised WHO Classification and Treatment of Childhood Pneumonia at Health Facilities: Evidence Summaries. World Health Organization. 2014; 26 p.

8. Report of a consultative meeting of children with pneumonia and HIV infection.

9. Short S, Bashir H, Marshall P, Miller N, Olmschenk D, Prigge K, Solyntjes L. Diagnosis and Treatment of Respiratory Illness in Children and Adults. Inst Clin Syst Improv. 2017; www.icsi.org

10. Rice, AL; Sacco, L; Hyder, A; Black R. Malnutrition as an underlying cause of childhood deaths associated with infectious diseases in developing countries. Bull World Health Organ. 2000;78(10):1207–21.

11. Macintyre CR, Chughtai AA, Zhang Y, Seale H, Yang P, Chen J, Yang P, Zhang D, Wang Q. Viral and bacterial upper respiratory tract infection in hospital health care workers over time and association with symptoms. 2017;1–9.

12. Midzi N, Mduluza T, Chimbari MJ, Tshuma C, Charimari L, Mhlanga G, Manangazira P, Munyati SM, Phiri I, Mutambu SL, Midzi SS, Ncube A, Muranzi LP, Rusakaniko S, Mutapi F (2014) Distribution of Schistosomiasis and Soil Transmitted Helminthiasis in Zimbabwe: Towards a National Plan of Action for Control and Elimination. PLoS Negl Trop Dis 8(8): e3014. doi:10.1371/journal.pntd.0003014.

13. Nelwan ML. Schistosomiasis: Life Cycle, Diagnosis, and Control. Curr Ther Res - Clin Exp [Internet]. 2019;91(24):5–9. https://doi.org/10.1016/j.curtheres.2019.06.001

14. Osakunor DNM, Mduluza T, Midzi N, Chase-Topping M, Mutsaka-Makuvaza MJ, Chimponda T. Dynamics of paediatric urogenital schistosome infection, morbidity and treatment: A longitudinal study among preschool children in Zimbabwe. BMJ Glob Heal. 2018;3(2):1–9.

15. Jenipher M, Makuvaza M, Zingoni ZM, Katsidzira A, Tshuma C, Chin’ombe N, Zhou X, Webster B, Midzi N. Urogenital schistosomiasis and risk factors of infection in mothers and preschool children in an endemic district in Zimbabwe. Parasit Vectors. 2019;1–15.

16. History T. Zimbabwe Control of Schistosomiasis. 2016;2015–6.

17. Taylor P, Makura O. Prevalence and distribution of schistosomiasis in Zimbabwe. Ann Trop Med Parasitol. 1985;79(3):287–99.

18. Mutsaka-Makuvaza MJ, Matsena-Zingoni Z, Tshuma C, Ray S, Zhou XN, Webster B, Midzi N. Reinfection of urogenital schistosomiasis in pre-school children in a highly endemic district in Northern Zimbabwe□: a 12 months compliance study. Infect Dis Poverty. 2018;7(1):102. doi: 10.1186/s40249-018-0483-7. PMID: 30268157

19. Schwartz E. Pulmonary schistosomiasis. 2002;23:433–43.

20. Schaberg T, Rahn W, Racz P, Lode H. Pulmonary schistosomiasis resembling acute pulmonary tuberculosis. Eur Respir J. 1991;4(8):1023–6.

21. Ross G A,Vickers D, Olds R G SSDPm. katayama syndrome. lancet Infect Dis. 2007;7(3):218–24.

22. Case A, Fever K, Schistosomiasis A. A Case of Katayama Fever (Acute Schistosomiasis). Med J Armed Forces India. 2010;66(3):299.

23. Esserman LJ, Yau C, Thompson CK, Veer LJ Van, Borowsky AD, Hoadley KA, Tobin NP, Nordenskjöld B, Fornander T, Stå l O, Benz CC, Lindström LS. Use of Molecular Tools to Identify Patients With Indolent Breast Cancers With Ultralow Risk Over 2 Decades. 2017;3(11):1503–10.

24. Freer JB, Bourke CD, Durhuus GH, Kjetland EF, Prendergast AJ. Review Schistosomiasis in the first 1000 days. Lancet Infect Dis. 2018;18(6):e193–203. http://dx.doi.org/10.1016/S1473-3099(17)30490-5

25. World Health Organization. Schistosomiasis Key facts. World Heal Organ. 2018;1–5. https://www.who.int/en/news-room/fact-sheets/detail/schistosomiasis

26. WHO. schistosomiasis. Global Health Estimates 2015: Deaths by Cause, Age, Sex, by Country and by Region, 2000-2015. 2016. http://www.who.int/healthinfo/global_burden_disease/estimates/en/index1.html%0AGeneva, World Health Organization; 2016.

27. Zimbabwe National Statistic Agency. Provincial report: Mashonaland Central. Zimbabwe Popul Census. 2012;

28. Osakunor DNM, Mduluza T, Midzi N, Chase-Topping M, Mutsaka-Makuvaza MJ, Chimponda T, Eyoh E, Mduluza T, Pfavayi LT, Wami WM, Amanfo SA, Murray J, Tshuma C, Woolhouse MEJ, Mutapi F. Dynamics of paediatric urogenital schistosome infection, morbidity and treatment: a longitudinal study among preschool children in Zimbabwe. BMJ Glob Health. 2018 ;3(2):e000661. doi: 10.1136/bmjgh-2017-000661. PMID: 29616147

29. Carter J, Müller-Stöver I, Östensen H. Good clinical diagnostic practice A guide for clinicians in developing countries to the clinical diagnosis of disease and to making proper use of clinical diagnostic services. 2005.

30. University of Glasgow. MB ChB Clinical History and Examination Manual. 2015;

31. Child TTHE. IMCI chart Booklet. 2002;1–39.

32. Tantilipikorn P, Auewarakul P. Airway allergy and viral infection. Asian Pacific J Allergy Immunol. 2011;29(2):113–9.

33. Center for Disease Control and Prevention. Nonspecific Upper Respiratory Tract Infection. Cent Dis Control Prev. 2006;134(6):2001.

34. World Health Organization. Recommendations for management of common childhood conditions. Parassitologia. 2012;84. http://europepmc.org/abstract/MED/10697870

35. Mott, K., Baltes, R., Bambagha J. and Baldassini B. Field studies for detection of a reusable polyamide filter for detection of Schistosoma haematobium eggs by urine filtration. ropenmedizin Parasitol 1982;33227-228. 1982;(33):227–8.

36. Wami WM, Nausch N, Bauer K, Midzi N, Gwisai R, Simmonds P, Mduluza T, Woolhouse M, Mutapi F. Comparing parasitological vs serological determination of Schistosoma haematobium infection prevalence in preschool and primary school-aged children: Implications for control programmes. Parasitology. 2014;141(14):1962–70.

37. Risk A, Risk PA. Appendix III-B. :155–62.

38. Risk PA. Definition of PAR PAR = Population Attributable Risk. 2014;8–11.

39. Lambertucci JR. Acute schistosomiasis mansoni: Revisited and reconsidered. Mem Inst Oswaldo Cruz. 2010;105(4):422–35.

40. Town C, Africa S, Saade A, Carton E, Damotte D, Yera H. Lung Involvement in Chronic Schistosomiasis with Bladder Squamous Cell Carcinoma. 2018;24(86949):2375–8.

41. King CH, Galvani AP. Underestimation of the global burden of schistosomiasis. Lancet. 2018;391(10118):307–8. http://dx.doi.org/10.1016/S0140-6736(18)30098-9

42. Kolosionek E, Graham BB, Tuder RM, Butrous G. Pulmonary vascular disease associated with parasitic infection — the role of schistosomiasis. Clin Microbiol Infect. 2010;17(1):15–24.: http://dx.doi.org/10.1111/j.1469-0691.2010.03308.x

43. Mazigo HD, Kirway L, Ambrose EA. Prevalence and intensity of Schistosoma mansoni infection in pediatric populations on antiretroviral therapy in north-western Tanzania: A cross-sectional study. BMJ Open. 2019;9(7):1–6.

44. Mott KE, Dixon H, Osei-Tutu E, England EC. Relation between Intensity of Schistosoma haematobium Infection and Clinical Haematuria and Proteinuria. Lancet. 1983;321(8332):1005–8.: https://doi.org/10.1016/S0140-6736(83)92641-7

45. Chipeta M, Ngwira B, Kazembe L. Analysis of Schistosomiasis haematobium Infection Prevalence and Intensity in Chikhwawa, Malawi: An Application of a Two Part Model. PLoS Negl Trop Dis. 2013;7:e2131.

46. Lucia C, Silva M. Endothelial Cells as Targets of the Intravascular Parasitic Disease Schistosomiasis a. Vascular Responses to Pathogens. Elsevier Inc.; 2016. 195–207 p.: http://dx.doi.org/10.1016/B978-0-12-801078-5/00015-7

47. Vennervald BJ, Dunne DW. Morbidity in schistosomiasis: An update. Curr Opin Infect Dis. 2004;17(5):439–47.

48. Weerakoon KGAD, Gobert GN, Cai P, Mcmanus P. Advances in the Diagnosis of Human Schistosomiasis. 2015;28(4):939–67.

49. Dondo V, Mujuru H, Nathoo K, Jacha V, Tapfumanei O, Chirisa P, Manangazira P, Macharaga J, de Gouveia L, Mwenda JM, Katsande R, Weldegebriel G, Pondo T, Matanock A, Lessa. Pneumococcal Conjugate Vaccine Impact on Meningitis and Pneumonia Among Children Aged < 5 Years - Zimbabwe, 2010 – 2016. 2019;69:2010–6.

